# Feasibility and Utility of a Structured Guide for Cannabis Tolerance Breaks in Young Adults

**DOI:** 10.1101/2022.05.18.22274748

**Authors:** Thomas JK Fontana, Jonathan A. Schulz, Alan J. Budney, Andrea C. Villanti

**Author notes:** **Corresponding Author:** Thomas Fontana, MS, 112 Davis Center, 590 Main Street, Burlington, VT 05401. **Financial Disclosure: TF** is the owner of Cum Laude Consulting LLC, which received a grant from the State of Vermont to investigate the T-Break Guide. Those funds were then subcontracted to the University of Vermont for this research.

## Abstract

**Objective:** To explore the feasibility and utility of a tolerance break (T-Break) guide on young adults’ cannabis use.

**Participants:** Young adults aged 18-29 (n=125) who were current cannabis users.

**Methods:** Participants recruited through posters and listservs at various universities were offered the T-Break Guide—daily activities, advice, and encouragement—to help complete a 21-day cannabis break. Bivariate analyses examined associations between Guide use and follow-up measures.

**Results:** Compared to non-Guide users, participants who used the Guide “a lot” were more likely to complete the 21-day abstinence break (84% vs. 57%), revise their personal definition of balance to mean less cannabis (84% vs. 62%), and plan a future break (32% vs. 11%).

**Conclusion:** Use of this self-directed Guide may help young adults take a break from cannabis use and reduce future use. Further research using more rigorous designs to test the Guide’s efficacy and assess longer-term maintenance of effects is needed.

## INTRODUCTION

In 2021, approximately 40% of undergraduate students reported using cannabis at least monthly and 14.6% reported using cannabis daily or almost daily.^1^ Cannabis use can negatively affect academic outcomes in college students, with cannabis-using students being more likely to drop out of college, have a lower grade point average, and miss classes than students who do not use cannabis.^2,3^ Cannabis use can also have negative effects on attention, memory, and processing speed.^4-6^

Effective interventions addressing cannabis use disorders include contingency management, cognitive-behavior therapy, and motivational enhancement.^7-10^ These interventions have consistently produced improvements in abstinence or reduction in use.^8-10^ However, these interventions can be resource intensive, requiring monetary commodities for incentives,^11^ or specialized therapists to implement.

Brief interventions to address cannabis use in nontreatment-seeking adolescents (e.g., brief in-person or phone sessions, ecological momentary assessment interviews) have resulted in reduced cannabis use, fewer cannabis problems, and fewer cannabis use disorder symptoms, maintained at one year.^7,12-15^ Although brief interventions have demonstrated initial positive results, effects have been small.^7,16^ Moreover, cannabis abstinence may not be the primary goal for young adults, consistent with low desire to quit cannabis in this population.^17^ Cannabis tolerance breaks, or T-breaks, have gained popularity online as a way to use cannabis responsibly, as suggested by posts within a sub-Reddit community with over 99,000 adult members.^18^ A T-break generally involves a self-directed process of abstaining from cannabis for a predetermined amount of time, with the aim of using less cannabis after the break and developing a balance in cannabis use. A break from cannabis use may also comprise a “practice quit attempt” identified in tobacco literature as a key pre-cessation target to increase coping skills and self-efficacy for future quit success.^19-21^ While there is little research on T-breaks for reducing cannabis use and other cannabis-related outcomes, they may offer a novel and acceptable approach to reducing cannabis use in nontreatment-seeking young adults.

This pilot study is the first to evaluate the feasibility and utility of a T-Break Guide with young adult cannabis users. The Guide comprises prescribed daily activities, advice, and encouragement designed to help young adults complete a break from cannabis. This study evaluated completion of the break, worth of the break, definition of balance, future plans to take a break, cannabis use after the break, and changes in motivation, confidence, and other drug and alcohol use in a small sample of young adult cannabis users.

## METHODS

### Participants and Procedures

The pilot study used a pretest–posttest design in which participants completed a baseline survey and were asked to attempt a 21-day period of cannabis abstinence, to consider using the T-Break Guide (intervention) in their attempt, and to complete a follow-up survey after 30 days. Participants were young adults aged 18-29 recruited through posters and tabling events at the University of Vermont (UVM), a post on a UVM webpage, emails on listservs of seven other universities, and a post on a subreddit focusing on responsible cannabis use. The posters and online announcements directed participants to the online baseline survey, which contained consent information before allowing participants to complete. After completing the baseline survey, participants received a link to an online version of the T-Break Guide through email. After 30 days, participants received another email prompting them to complete the follow-up survey. Participants completed both surveys online using Impact Feedback software and were compensated $10 for completing the baseline survey and $20 for completing the follow-up survey, regardless of whether they used the T-Break Guide or started a break. The UVM Institutional Review Board reviewed and approved all study procedures (STUDY00001489).

A total of 151 people completed the baseline survey and 143 (95%) completed both baseline and follow-up surveys. A subset (n=125) were included in analyses after excluding those who did not meet the age criterion (ages 18– 29; n = 14), did not reply to use of the Guide (n = 1), or indicated no current cannabis use (n = 3).

### Intervention

The T-Break Guide was designed to help people complete a 21-day break from cannabis. For each day of the break, the Guide offers inspiration in the form of a quote, reflections based on likely experiences occurring at that point during the break, advice on ways to overcome challenges, alternative activities in which to engage, and encouragement. The first week focuses on physical symptoms of cannabis withdrawal (e.g., sleep, appetite); the second week focuses on the emotional experience (e.g., anxiety, boredom); and the third week focuses on behavioral aspects (e.g., examining patterns, connections). The T-Break Guide follows the Motivational Interviewing principles of affirmation and autonomy.^22^

### Measures

#### Baseline Measures

Participants provided sociodemographic data on age, gender, race/ethnicity, sexual orientation, physical disability, mental illness diagnosis, educational status, employment status, other drug use, ‘medical card’ status, frequency of cannabis use, motivation to use cannabis, enjoyment of cannabis use, and age of first cannabis use at baseline.

#### Use of the T-Break Guide

At follow-up, participants were asked, “You were asked to consider using the T-Break Guide. Did you use it?” with four response options: “No, I never looked,” “Not really, I looked but it didn’t appeal to me,” “I used it some,” and “I used it a lot.” “No, I never looked” and “not really, I looked but it didn’t appeal to me” were collapsed into one category as participants who did not use the Guide. Three resulting categories reflected use of the Guide: “No use”, “Some use,” and “A lot of use.”

#### Cannabis and Other Substance Use

At follow-up, participants responded to a question about whether they completed a 21-day break from cannabis. If not, they were asked if they completed their own pre-determined goal. All participants responded to questions asking if taking a tolerance break was worth it, about their cannabis use after completing a break, their confidence in taking a future break, change in their personal definition of “balance” around cannabis, the importance of finding/keeping balance with cannabis, where they picture their use three years from now, if they plan to take a break again, and about their alcohol and other drug use during their T-break.

Additionally, participants responded to open-ended questions about their experience during the study, including what (if any) aspects of the T-Break Guide they found useful, if the break allowed them to do anything that would have been more difficult if they were still consuming cannabis, and why they continued the break, if they did.

#### Data Analysis

All analyses were run using Stata/Standard edition version 17 (StataCorp LP, College Station, TX). Missing data were deleted using list-wise deletion per Stata’s default procedures. We ran descriptive statistics for the sociodemographic variables and tested each variable for differences in distribution between those who did or did not use the T-Break Guide using chi-squared tests. Bivariate analyses using chi-squared tests assessed differences in each follow-up measure by participants’ use of the Guide status (no use, some use, a lot of use).

## Results

Participants were 22.1 years on average and the majority were male, white, currently enrolled as undergraduates, and reported cannabis use 4 or more times a week. Approximately one-third of participants had a mental illness diagnosis (Table S1). Of the 125 participants, 37% did not use the T-Break Guide, 38% used the Guide “some,” and 25% used the Guide “a lot.” Table 1 displays responses to follow-up questions by T-Break Guide use. Overall, 77 (62%) participants successfully completed the T-break, and of those who did not complete the break an additional six (13%) successfully completed their personal cannabis abstinence goal. Compared to those who did not use the Guide or used the Guide “some,” participants who used the T-Break Guide “a lot” were more likely to complete a 21-day tolerance break (84% vs. 54%; p=0.003), plan a future break (32% vs. 10%; p=0.011), report that their personal definition of balance had evolved to mean less cannabis (84% vs. 55%; p=0.03), and report that they do not see themselves using cannabis in three years (36% vs 4%; p<0.0001). A greater proportion of Guide users reported that the T-break was worth it than Guide non-users (89% vs. 53%; p=0.008). Finally, among those who reported alcohol use, people who completed the T-break were more likely to report stopping or drinking less during the break than those who did not complete the T-break (67% vs 27%; p=0.026). Among those who reported other drug use, those who completed the T-break were more likely to report stopping using other drugs or using less other drugs during the break (77% vs. 36%; p=0.042).

Components of the T-Break Guide that participants reported in the follow-up survey finding useful included the quotes, activities/resources, daily progress checks, and the validation it provided. When asked what they could now accomplish because they were not using cannabis, participants reported they were able to improve their finances, repair relationships with family and friends, focus on their studies, concentrate better, and exercise more. However, some participants noted that stopping cannabis use did not allow them to do anything that would have been more difficult if they were still using cannabis. Responses to why a participant was able to continue their break after successfully completing their break included no longer having a desire to use cannabis, feeling better, being busy, or having an upcoming drug test. For those who reported not using the Guide, reasons included forgetting about it or not needing it as they were already motivated.

## DISCUSSION

Overall, 62% of young adults in this initial study of the T-Break Guide successfully completed a 21-day break from cannabis, and an additional 12% of the participants stopped using cannabis in line with their predetermined goal. A greater proportion of those who used the Guide “a lot” compared to those who used the Guide “some” or “no/not really” reported T-break completion, plans for a future T-break, and a revised personal definition of balance to mean less cannabis use. Ten days after the break period ended, 48% of participants who completed the T-break continued to refrain from cannabis use. Completing a T-break did not appear to increase alcohol and other drug use in most participants.

Prior studies have highlighted the importance of a voluntary check-up for cannabis use in young people,^12,13^ but there was no empirical evidence on the utility of a voluntary break from cannabis, nor the feasibility of this approach to reducing young adult cannabis use. Our findings suggest that the use of this low touch, self-directed guide may help young adults abstain from cannabis and that completing a T-break may increase confidence in future break success, re-evaluation of cannabis use, and plans for a future break. Our data also suggest that use of the Guide and abstinence from cannabis did not lead to increased alcohol use in our sample, which has been raised as a concern in prior investigations with young adults who discontinue cannabis use.^23^

Treatments for cannabis use disorders, even brief interventions, can be resource intensive. Our initial findings suggest that provision of a novel, low-burden, self-directed online guide provides an approach that may effectively encourage young adults to reduce their cannabis use. This type of self-directed guide may have a wide reach and is potentially scalable as resources needs are minimal. Additionally, this intervention may be consistent with some individual’s personal preference for responsible cannabis use rather than complete abstinence and also engage participants who were not actively seeking treatment.

There are a number of limitations to this initial pilot study. First, the participants self-reported cannabis use and Guide use; no methods were used to validate self-reports. Second, participants self-selected into the study and therefore may represent a sample of those who use cannabis who are particularly motivated to take a cannabis break. Last, our study did not include a control group and the sample size was small. These limitations are consistent with the pilot nature of this study. Future, more rigorous research, should evaluate the utility of cannabis tolerance breaks as an entry into reducing cannabis use and the efficacy of practical resources, like the T-Break Guide, in cannabis cessation efforts for young adults.

## Data Availability

All data produced are available online at https://doi.org/10.7910/DVN/RQZQ1Q

## References

1. American College Health Association. American College Health Association-National College Health Assessment III: Undergraduate Student Reference Group Data Report Spring 2021. Silver Spring, MD: American College Health Association; 2021

2. Suerken CK, Reboussin BA, Egan KL, et al. Marijuana use trajectories and academic outcomes among college students. Drug Alcohol Depend. 2016;162:137–145. doi:10.1016/j.drugalcdep.2016.02.041

3. Arria AM, Caldeira KM, Bugbee BA, Vincent KB, O’Grady KE. The academic consequences of marijuana use during college. Psychol Addict Behav. 2015;29(3):564–575. doi:10.1037/adb0000108

4. Pope HG, Yurgelen-Todd D.. The residual cognitive effects of heavy marijuana use in college students. JAMA. 1996;275(7):521–527. doi:10.1001/jama.275.7.521

5. Bolla KI, Brown K, Eldreth D, Tate K, Cadet JL. Dose-related neurocognitive effects of marijuana use. Neurology. 2002;59(9):1337–1343. doi:10.1212/01.WNL.0000031422.66442.49

6. Croft RJ, Mackay AJ, Mills ATD, Gruzelier JGH. The relative contributions of ecstasy and cannabis to cognitive impairment. Psychopharmacology. 2001;153(3):373–379. doi:10.1007/s002130000591

7. Winters KC, Mader J, Budney AJ, Stanger C, Knapp AA, Walker DD. Interventions for cannabis use disorder. Curr Opin Psychol. 2021;38:67–74. doi:10.1016/j.copsyc.2020.11.002

8. Litt MD, Kadden RM, Petry NM. Behavioral treatment for marijuana dependence: Randomized trial of contingency management and self-efficacy enhancement. Addict Behav. 2013;38(3):1764–1775. doi:10.1016/j.addbeh.2012.08.011

9. Budney AJ, Fearer S, Walker DD, et al. An initial trial of a computerized behavioral intervention for cannabis use disorder. Drug Alcohol Depend. 2011;115(1-2):74–79. doi:10.1016/j.drugalcdep.2010.10.014

10. Banes KE, Stephens RS, Blevins CE, Walker DD, Roffman RA. Changing motives for use: Outcomes from a cognitive-behavioral intervention for marijuana-dependent adults. Drug Alcohol Depend. 2014;139:41–46. doi:10.1016/j.drugalcdep.2014.02.706

11. Schuster RM, Hanly A, Gilman J, Budney A, Vandrey R, Evins AE. A contingency management method for 30-days abstinence in non-treatment seeking young adult cannabis users. Drug Alcohol Depend.. 2016;167:199–206. doi:10.1016/j.drugalcdep.2016.08.622

12. Walker DD, Stephens R, Roffman R, et al. Randomized controlled trial of motivational enhancement therapy with nontreatment-seeking adolescent cannabis users: A further test of the teen marijuana check-up. Psychol Addict Behav. 2011;25(3):474–484. doi:10.1037/a0024076

13. Walker DD, Stephens RS, Blevins CE, Banes KE, Matthews L, Roffman RA. Augmenting brief interventions for adolescent marijuana users: The impact of motivational check-ins.J Consult Clin Psychol. 2016;84(11):983–992. doi:http://dx.doi.org/10.1037/ccp0000094

14. Prince MA, Collins RL, Wilson SD, Vincent PC. A preliminary test of a brief intervention to lessen young adults’ cannabis use: Episode-level smartphone data highlights the role of protective behavioral strategies and exercise. Exp Clin Psychopharmacol. 2020;28(2):150–156. doi:10.1037/pha0000301

15. Swan M, Schwartz S, Berg B, Walker D, Stephens R, Roffman R. The Teen Marijuana Check-Up: An in-school protocol for eliciting voluntary self-assessment of marijuana use. J Soc Work Pract Addict. 2008;8(3):284–302. doi:10.1080/15332560802223305

16. Halladay J, Scherer J, MacKillop J, et al. Brief interventions for cannabis use in emerging adults: A systematic review, meta-analysis, and evidence map. Drug Alcohol Depend. 2019;204:107565–107565. doi:10.1016/j.drugalcdep.2019.107565

17. Ramo DE, Liu H, Prochaska JJ. Reliability and validity of young adults’ anonymous online reports of marijuana use and thoughts about use. Psychol Addict Behav. 2012;26(4):801–811. doi:10.1037/a0026201

18. r/Petrioles – For the reduction, moderation and responsible consumption of cannabis. Reddit. https://www.reddit.com/r/Petioles/. Accessed November 10, 2021.

19. Baker TB, Mermelstein R, Collins LM, et al. New methods for tobacco dependence treatment research. Ann Behav Med. 2011;41(2):192–207. doi:10.1007/s12160-010-9252-y

20. Practice quitting programs. smokefree.gov. https://smokefree.gov/tools-tips/text-programs/practice-quitting. Accessed November 10, 2021.

21. Every try counts campaign. U.S. Food & Drug Administration. https://www.fda.gov/tobacco-products/every-try-counts-campaign. Accessed November 10, 2021

22. Hall K, Gibbie T, Lubman DI. Motivational interviewing techniques – facilitating behavior change in the general practice setting. Aust Fam Physician. 2012;14(9):660–667

23. Schuster RM, Potter K, Lamberth E, et al. Alcohol substitution during one month of cannabis abstinence among non-treatment seeking youth. Prog Neuropsychoparmacol Biol Psychiatry. 2021;107:110205. doi:https://doi.org/10.1016/j.pnpbp.2020.110205

