## Supplementary material for "Feasibility and Utility of a Structured Guide for Cannabis Tolerance Breaks in Young Adults": Table - 1 of 1

*Table 1 Participant responses at follow-up, by use of the T-Break Guide (n = 125)*

|  |  | T-Break Guide User | | | |  |  |
| --- | --- | --- | --- | --- | --- | --- | --- |
|  |  | No (n=46)  n, % | Some (n=48)  n, % | | A lot (n=31)  n, % | Total (n=125)  n, % | *p-*value |
| Did you complete a 21-day tolerance break? | | | | | | | .012 |
|  | No | 20 (43.5) | 23 (47.9) | | 5 (16.1) | 48 (38.4) |  |
|  | Yes | 26 (56.5) | 25 (52.1) | | 26 (83.9) | 77 (61.6) |  |
| **Participants who completed the T-Break** | | | | | | |  |
| Was the T-Break worth it? | | | | | | | .011 |
|  | Not Really | 2 (7.7) | 0 (0) | | 0 (0) | 2 (2.6) |  |
|  | Mostly | 12 (46.2) | 10 (40.0) | | 3 (11.5) | 25 (32.5) |  |
|  | Yes | 12 (46.2) | 15 (60.0) | | 23 (88.5) | 50 (64.9) |  |
| You went the 21 days without  cannabis, then what? | |  | | | | | .631 |
|  | I got high immediately | 3 (11.5) | 3 (12.0) | | 2 (7.7) | 8 (10.4) |  |
|  | I got high eventually | 10 (38.5) | 13 (52.0) | | 9 (34.6) | 32 (41.6) |  |
|  | I kept the break going | 13 (50.0) | 9 (36.0) | | 15 (57.7) | 37 (48.1) |  |
| **Participants who completed T-break and then got high immediately or eventually** | |  | | | | |  |
| After break, about how much did it take  to get you high (compared to same high  just before your break)? | |  | | | | | .045 |
|  | Less than ½ | 8 (61.5) | 2 (12.5) | | 5 (45.5) | 15 (7.0) |  |
|  | About ½ | 4 (30.8) | 10 (62.5) | | 3 (27.3) | 17 (7.8) |  |
|  | About ¾ | 0 (0) | 4 (25.0) | | 3 (27.3) | 7 (62.8) |  |
|  | Pretty much the same | 1 (7.7) | 0 (0) | | 0 (0) | 1 (14.7) |  |
| In the days since your break ended, how does your cannabis use compare to before break? | |  |  | |  |  | 0.111 |
|  | Way less | 7 (53.8) | 4 (25.0) | | 7 (63.6) | 18 (45.0) |  |
|  | A little less | 5 (38.5) | 6 (37.5) | | 3 (27.3) | 14 (35.0) |  |
|  | Same | 1 (7.7) | 1 (6.3) | | 1 (9.1) | 3 (7.5) |  |
|  | A little more | 0 (0.0) | 5 (31.3) | | 0 (0.0) | 5 (12.5) |  |
|  | Way more | 0 (0.0) | 0 (0.0) | | 0 (0.0) | 0 (0.0) |  |
| **Participants who did not complete T-Break** | |  |  | |  |  |  |
| If not completed, goal complete? | | | | | | | .261 |
|  | No | 16 (80.0) | 22 (95.7) | | 4 (80.0) | 42 (87.5) |  |
|  | Yes | 4 (20.0) | 1 (4.3) | | 1 (20.0) | 6 (12.5) |  |
| **All Participants** | |  | | | | |  |
| Will you take a break again? | |  | | | | | .024 |
|  | No | 0 (0.0) | 1 (2.1) | | 0 (0.0) | 1 (0.8) |  |
|  | Not likely | 5 (10.9) | 2 (4.2) | | 0 (0.0) | 7 (5.6) |  |
|  | Yes, likely | 36 (78.3) | 41 (85.4) | | 21 (67.7) | 98 (78.4) |  |
|  | Yes, and I already have it planned | 5 (10.9) | 4 (8.3) | | 10 (32.3) | 19 (15.2) |  |
| Where do you picture your cannabis use in 3 years? I picture using: | |  | | | | | <.0001 |
|  | None | 2 (4.3) | 2 (4.2) | | 11 (35.5) | 15 (12.0) |  |
|  | A lot less | 4 (8.7) | 18 (37.5) | | 11 (35.5) | 33 (26.4) |  |
|  | A little less | 21 (45.7) | 19 (39.6) | | 5 (16.1) | 45 (36.0) |  |
|  | Same | 17 (37.0) | 4 (8.3) | | 3 (9.7) | 24 (19.2) |  |
|  | A little more | 2 (4.3) | 3 (6.3) | | 1 (3.2) | 6 (4.8) |  |
|  | A lot more | 0 (0) | 2 (4.2) | | 0 (0) | 2 (1.6) |  |
| How has your personal definition of balance changed in the past month? | |  | | | | | 0.177 |
|  | It has evolved to mean less cannabis | 25 (54.3) | | 27 (56.3) | 26 (83.9) | 78 (62.4) |  |
|  | It has not changed | 19 (41.3) | | 18 (37.5) | 5 (16.1) | 42 (33.6) |  |
|  | It has evolved to mean more cannabis | 1 (2.2) | | 1 (2.1) | 0 (0.0) | 2 (1.6) |  |
|  | Other | 1 (3.3) | | 2 (4.2) | 0 (0.0) | 3 (2.4) |  |
| If you wanted to take a tolerance break in the future, how confident are you that you can be successful? (0=*not confident*, 10=*very confident*), mean score (SD) | |  |  | |  |  | <.0001 |
|  | Completed T-Break | 9.25 (1.1) | 8.9 (1.2) | | 8.9 (1.1) | 9.0 (1.1) |  |
|  | Did not complete T-Break | 7.3 (1.6) | 7.1 (1.6) | | 8.0 (1.9) | 7.3 (1.6) |  |
| How important is it to you to find/keep balance with cannabis? ((0=*not important*, 10=*very important*), mean score (SD) | |  |  | |  |  | .633 |
|  | Completed T-Break | 8.3 (2.0) | 8.25 (1.8) | | 9.1 (1.3) | 8.5 (1.7) |  |
|  | Did not complete T-Break | 8.5 (2.1) | 8.1 (2.3) | | 7.6 (1.7) | 8.2 (2.1) |  |
