## Supplemental Table - 1 of 1 for "Feasibility and Utility of a Structured Guide for Cannabis Tolerance Breaks in Young Adults"

*Table S1. Demographics of young adults enrolled in the T-Break Guide study (n = 125)*

|  |  | T-Break Guide User | | |  |
| --- | --- | --- | --- | --- | --- |
|  |  | No (n=46)  n, % | Some (n=48)  n, % | A lot (n=31)  n, % | Total (n=125)  n, % |
| Age, mean years (SD) | | 20.1 (1.7) | 22.9 (3.3) | 23.7 (2.9) | 22.1 (3.1) |
| Gender^a^ |  | | | | |
|  | Trans | 2 (4.3) | 3 (6.3) | 3 (9.7) | 8 (6.4) |
|  | Female | 24 (52.2) | 13 (27.1) | 11 (35.5) | 48 (38.4) |
|  | Male | 20 (43.5) | 32 (66.7) | 17 (54.8) | 69 (55.2) |
| Race/Ethnicity^a^ | | | | | |
|  | Non-white | 7 (15.2) | 13 (27.1) | 9 (29.0) | 29 (23.2) |
|  | White | 39 (84.8) | 35 (72.9) | 22 (71.0) | 96 (76.8) |
| Sexual Orientation^a^ | | | | | |
|  | blank/questioning | 3 (6.5) | 4 (8.3) | 0 (0) | 7 (5.6) |
|  | gay/queer | 5 (10.9) | 4 (8.3) | 4 (12.9) | 13 (10.4) |
|  | bi/pan | 11 (23.9) | 6 (12.5) | 5 (16.1) | 22 (17.6) |
|  | straight | 27 (58.7) | 34 (70.8) | 22 (71.0) | 83 (66.4) |
| Physical Disability | | | | | |
|  | No | 45 (97.8) | 48 (100) | 31 (100) | 124 (99.2) |
|  | Yes | 1 (2.2) | 0 (0) | 0 (0) | 1 (0.8) |
| Ever Diagnosed with Mental Illness | | | | | |
|  | No | 25 (54.3) | 35 (72.9) | 24 (77.4) | 84 (67.2) |
|  | Yes | 21 (45.7) | 13 (27.1) | 7 (22.6) | 41 (32.8) |
| Educational Status | | | | | |
|  | No college | 0 (0) | 3 (6.3) | 6 (19.4) | 9 (7.2) |
|  | Did not complete college | 2 (4.3) | 4 (8.3) | 4 (12.9) | 10 (8.0) |
|  | Currently enrolled undergraduate | 41 (89.1) | 24 (50.0) | 12 (38.7) | 77 (61.6) |
|  | Bachelor’s or more | 3 (6.5) | 17 (35.4) | 9 (29.0) | 29 (23.2) |
| Employment Status | | | | | |
|  | Unemployed | 20 (43.5) | 14 (29.2) | 10 (32.3) | 44 (35.2) |
|  | Parttime | 21 (45.7) | 28 (58.3) | 9 (29.0) | 58 (46.4) |
|  | Fulltime | 5 (10.9) | 6 (12.5) | 12 (38.7) | 23 (18.4) |
| How often do you use cannabis? | |  |  |  |  |
|  | Monthly or less | 0(0) | 3 (6.3) | 1 (3.2) | 4 (3.2) |
|  | 2–4 times a month | 7 (15.6) | 1 (2.1) | 0 (0) | 8 (6.5) |
|  | 2–3 times a week | 13 (28.9) | 5 (10.4) | 4 (12.9) | 22 (17.7) |
|  | 4 or more times a week | 25 (55.6) | 39 (81.3) | 26 (83.9) | 90 (72.6) |
| Top motivation to use | |  |  |  |  |
|  | Medicinal | 2 (4.5) | 7 (14.6) | 6 (19.4) | 15 (12.2) |
|  | Inspiration/Creative | 5 (11.4) | 7 (14.6) | 5 (16.1) | 17 (13.8) |
|  | Self-medication | 7 (15.9) | 10 (20.8) | 4 (12.9) | 21 (17.1) |
|  | Spiritual/Meditative | 0 (0) | 1 (2.1) | 5 (16.1) | 6 (4.9) |
|  | Reward | 8 (18.2) | 6 (12.5) | 3 (9.7) | 17 (13.8) |
|  | Recreational | 22 (50) | 17 (35.4) | 8 (25.8) | 47 (38.2) |
| Age of first cannabis use, mean years (SD) | | 16.5 (1.5) | 17.7 (2.6) | 19.0 (2.7) | 17.6 (2.5) |

^a^ Gender, race/ethnicity, and sexual orientation were open-ended questions. We coded gender as “trans,” “female,” and “male;” race/ethnicity as “non-white” and “white;” and sexual orientation as “blank/questioning,” “gay/queer,” “bi/pan,” and “straight.”
